# Rates of regional tau accumulation in ageing and across the Alzheimer’s disease continuum: An AIBL ^18^F-MK6240 PET study

**DOI:** 10.1101/2022.03.11.22272240

**Authors:** Natasha Krishnadas, Vincent Doré, Jo Robertson, Larry Ward, Christopher Fowler, Colin L Masters, Pierrick Bourgeat, Jurgen Fripp, Victor L Villemagne, Christopher C Rowe

**Affiliations:** Florey Department of Neurosciences & Mental Health, The University of Melbourne, Parkville, VIC 3052, Australia; Department of Molecular Imaging & Therapy, Austin Health, Heidelberg, VIC 3084, Australia; Health and Biosecurity Flagship, The Australian eHealth Research Centre, Melbourne, Victoria, Australia; Florey Institute of Neurosciences & Mental Health, Parkville, VIC 3010, Australia; Health and Biosecurity Flagship, The Australian eHealth Research Centre, Brisbane, QLD, Australia; Department of Psychiatry, University of Pittsburgh, Pittsburgh, PA, USA

**Keywords:** Tau, ^18^F-MK6240, Positron Emission Tomography (PET), Alzheimer’s disease (AD), Longitudinal

## Abstract

**Background:** Tau PET imaging enables prospective longitudinal observation of the rate and location of tau accumulation in Alzheimer’s disease (AD). ^18^F-MK6240 is a newer, high affinity tracer for the paired helical filaments of tau in AD. It is widely used in clinical trials, despite sparse longitudinal natural history data. We aimed to evaluate the impact of disease stage, and two reference regions on the magnitude and effect size of regional change.

**Methods:** One hundred and fifty-eight participants: 83 cognitively unimpaired (CU) Aβ-, 37 CU Aβ+, 19 mild cognitively impaired (MCI) Aβ+ and 19 AD Aβ+ had annual ^18^F-MK6240 PET for one or two years (mean 1.6 years). Standardized uptake value ratios (SUVR) were generated for three in-house composite ROI: mesial temporal (Me), temporoparietal (Te), and rest of neocortex (R), and a Free-Surfer derived meta-temporal (MT) ROI. Two reference regions were examined: cerebellar cortex (SUVR_Cb_) and eroded subcortical white matter (SUVR_WM_).

**Results:** Low rates of accumulation were seen in CU Aβ-, predominantly in the mesial temporal lobe (MTL). In CU Aβ+, increase was greatest in the MTL, particularly the amygdala. In MCI Aβ+, a similar increase was seen in MTL, but also globally in the cortex. In AD Aβ+, greatest increase was in temporoparietal and frontal regions, with a decrease in the MTL. In CU and MCI increases were greater using SUVR_WM_. In AD, the SUVR_Cb_ showed marginally greater increase. Interpolation of the data suggests it takes approximately two decades to accumulate tau to the typical levels found in AD, similar to the rates of accumulation of Aβ plaques.

**Conclusions:** The rate of tau accumulation varies according to brain region and baseline disease stage, confirming previous reports. The PET measured change is greater, with fewer outliers, using an eroded white matter reference region, except in AD. While the eroded subcortical white matter reference may be preferred for trials in preclinical AD, the cerebellar cortex would be preferred for trials in symptomatic AD.

**Trial registration:** Not applicable.

## Background

Intracellular tau neurofibrillary tangles (NFT), along with extracellular amyloid-β (Aβ) plaques, are two pathological hallmarks of Alzheimer’s disease (AD). In AD, the spatial distribution of brain tau aggregates is linked with cognitive impairment in a domain-specific manner (1-3). Tau is also closely linked with the development of neurodegeneration (3-7). Longitudinal *in vivo* tracking of tau can facilitate our understanding of the natural trajectory of tau, its interactivity with Aβ, and associations with the clinical features of AD. Serial tau PET has also been proposed as a surrogate marker for disease progression in AD clinical trials (8) and has been included as an outcome measure in trial sub-analyses aiming to evaluate the impact of anti-Aβ therapies on tau burden (9, 10).

Tau tracer ^18^F-MK6240 has high affinity and selectivity for tau NFT in AD (11, 12). Serial imaging with ^18^F-MK6240 has reported detectable rates of tau accumulation in both preclinical and symptomatic AD stages (13). Detection of tau accumulation in preclinical AD is advantageous for clinical trials that are aiming to target this earlier stage of disease (14). The main aim of this study was to evaluate the rates of tau accumulation in ageing (CU Aβ-), preclinical AD (CU Aβ+), and symptomatic disease stages (Aβ+ mild cognitive impairment [MCI] and Alzheimer’s disease dementia [AD]) using ^18^F-MK6240. The second aim was to investigate the impact of two reference regions (cerebellar cortex and eroded subcortical white matter) on the magnitude and effect size of regional change.

## Methods

### Participants

Participants from the Australian Imaging Biomarkers and Lifestyle flagship study of ageing (AIBL) (15) and the Australian Dementia Network (ADNeT) who completed baseline and one or more follow-up tau ^18^F-MK6240 PET scans before July 2021 were included in this study if they met the following criteria: 1) ≥50 years of age; 2) were fluent in English; 3) had completed at least 7 years of education; and 4) did not have any history of neurological or psychiatric disorders, drug or alcohol abuse or dependence, or any other unstable medical condition. All participants complete neuropsychology assessments every 12 to 18 months, as previously described (15). Based on available clinical information and neuropsychology assessments, a multi-disciplinary clinical review panel, blind to Aβ and tau PET results, determines each participant’s clinical classification. Participants were deemed to be cognitively unimpaired (CU) if their performance on neuropsychology assessments was within 1.5 standard deviations of published normative data for their age group. A diagnosis of mild cognitive impairment (MCI) or Alzheimer’s disease dementia (AD) was assigned in accordance with international consensus criteria, as previously described (16). This study has been approved by the institutional review boards of all participating institutions, and all participants signed an informed consent form.

### Image acquisition

Tau PET imaging involved the intravenous administration of 185MBq (±10%) of ^18^F-MK6240 with a 20-minute acquisition commencing 90-minutes post-injection. Aβ PET imaging involved the intravenous administration of 200MBq (±10%) of ^18^F-NAV4694 with a 20-minute acquisition time commencing 50-minutes post-injection. PET scans were acquired on either a Philips TF64 PET/CT or a Siemens Biograph mCT. PET scans for each participant were performed on the same scanner at baseline and follow-up. Low dose CT was obtained for attenuation correction.

### Image analysis

Tau ^18^F-MK6240 PET scans were spatially normalized using the MR-less CapAIBL PCA-based approach (17) and scaled using two reference regions: 1) cerebellar cortex; and 2) eroded subcortical white matter. A gray matter inclusion mask and a meninges exclusion mask were applied. ^18^F-MK6240 standardized uptake value ratio (SUVR) was estimated for three in-house composite ROI: i) mesial temporal, Me (comprising entorhinal cortex, amygdala, hippocampus, and parahippocampus); ii) temporoparietal, Te (comprising inferior and middle temporal, fusiform, supramarginal and angular gyri, posterior cingulate/ precuneus, superior and inferior parietal, and lateral occipital cortex); and iii) rest of neocortex, R (comprising dorsolateral and ventrolateral prefrontal, orbitofrontal cortex, gyrus rectus, superior temporal and anterior cingulate), as previously described (18). Tau ^18^F-MK6240 SUVR was also estimated in a meta-temporal composite, MT (comprising Free-Surfer derived entorhinal cortex, parahippocampus, amygdala, inferior temporal, fusiform, and middle temporal cortex ROI) (19).

Aβ PET scans were spatially normalized using the MR-less CapAIBL approach (https://milxcloud.csiro.au/tools/capaibl), and the CapAIBL calibrated Centiloid method was applied for quantification (20). A Centiloid value of 25 was used to discriminate high (Aβ+) and low (Aβ-) Aβ scans.

### Vertex-based surface analysis

All ^18^F-MK6240 scans were in the same standard space and were projected onto the cortical surface using CapAIBL. Vertex-based surface analysis was used to estimate the mean annual rate of tau accumulation (change in SUVR/ year) for each of the clinical groups, scaled using two reference regions: 1) cerebellar cortex; and 2) eroded subcortical white matter.

### Statistical method

Demographic data were analyzed using an independent samples t-test for continuous data, and Chi-square test of independence for categorical data, with a threshold of significance of p<0.05. Effect size was reported as Cohen’s *d*. Annual change in ^18^F-MK6240 SUVR in each composite ROI was estimated as the slope obtained from linear regression of SUVR (independent variable) by Δtime (dependent variable). Annual percentage change in ^18^F-MK6240 SUVR in each composite was estimated as follows:

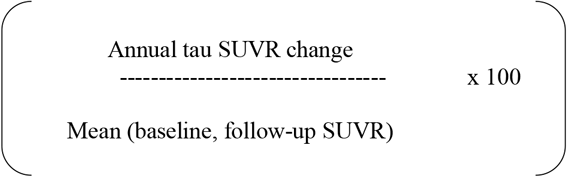

## Results

### Participants

The cohort comprised 158 participants: 83 CU Aβ-, 37 CU Aβ+, 38 cognitively impaired Aβ+ (19 with MCI and 19 with AD dementia). Table 1 shows the demographic characteristics of the cohort at baseline.

**Table 1.** Cohort demographic characteristics at baseline.

| | CU Aβ- | CU Aβ+ | MCI Aβ+ | AD Aβ+ |
| --- | --- | --- | --- | --- |
|  | *n* = 83 | *n* = 37 | *n* = 19 | *n* = 19 |
| Age (yrs) | 74.6±4.4 | 78.2±6.2** | 77.5±7.2 | 69.7±8.5* |
| Sex, F (%) | 48.2% | 56.8% | 47.4% | 42.1% |
| *APOE* ε4+ (%)<sup>a</sup> | 26.5% | 63.9%** | 52.9%* | 57.1%* |
| Centiloid | 4.5±9.1 | 80.1±38.5** | 99.9±51.3** | 117.7±38.0** |
| Baseline MMSE | 28.6±1.2 | 27.9±1.5* | 26.3±1.7** | 22.2±3.5** |
| Duration of follow-up (yrs) | 1.68 (0.47) | 1.73 (0.53) | 1.61 (0.49) | 1.50 (0.45) |
| T-test (one-tailed) * *p*<0.05, ** *p*<0.001 compared to CU Aβ-. | | | | |

<sup>a</sup> APOE genotype only available for 149/158 participants.

### Tau accumulation using vertex-based surface analysis

Vertex-based surface analysis was performed to demonstrate the spatial distribution and rate of ^18^F-MK6240 accumulation (SUVR/ year) across the clinical groups, normalized to the cerebellar cortex (Figure 1). Analysis in CU Aβ-individuals detected minimal tau accumulation. CU Aβ+ individuals showed tau accumulation in the mesial temporal cortex, inferior and lateral temporal gyri, and clusters in the precuneus and angular gyri. MCI Aβ+ individuals were observed to have tau accumulation in the precuneus, superior and lateral temporal, occipital, parietal, and prefrontal cortices. AD Aβ+ individuals were observed to have tau accumulation in the precuneus, temporal, parietal, occipital, and prefrontal cortices, with no accumulation in the mesial temporal cortex. The analysis was also performed using SUVR_WM_ (Supplementary Figure 1). With this reference region, CU Aβ-individuals were observed to have low rates of tau accumulation in the mesial temporal and temporal cortices. The spatial distribution of tau observed in the CU Aβ+, MCI Aβ+ and AD Aβ+ groups was similar, irrespective of the reference region. However, higher rates of tau accumulation were observed in the CU Aβ+ and MCI Aβ+ groups, and lower rates of tau accumulation were observed in the AD Aβ+ groups for SUVR_WM_ than SUVR_Cb_ (Figure 1, Supplementary Figure 1).

**Figure 1.**
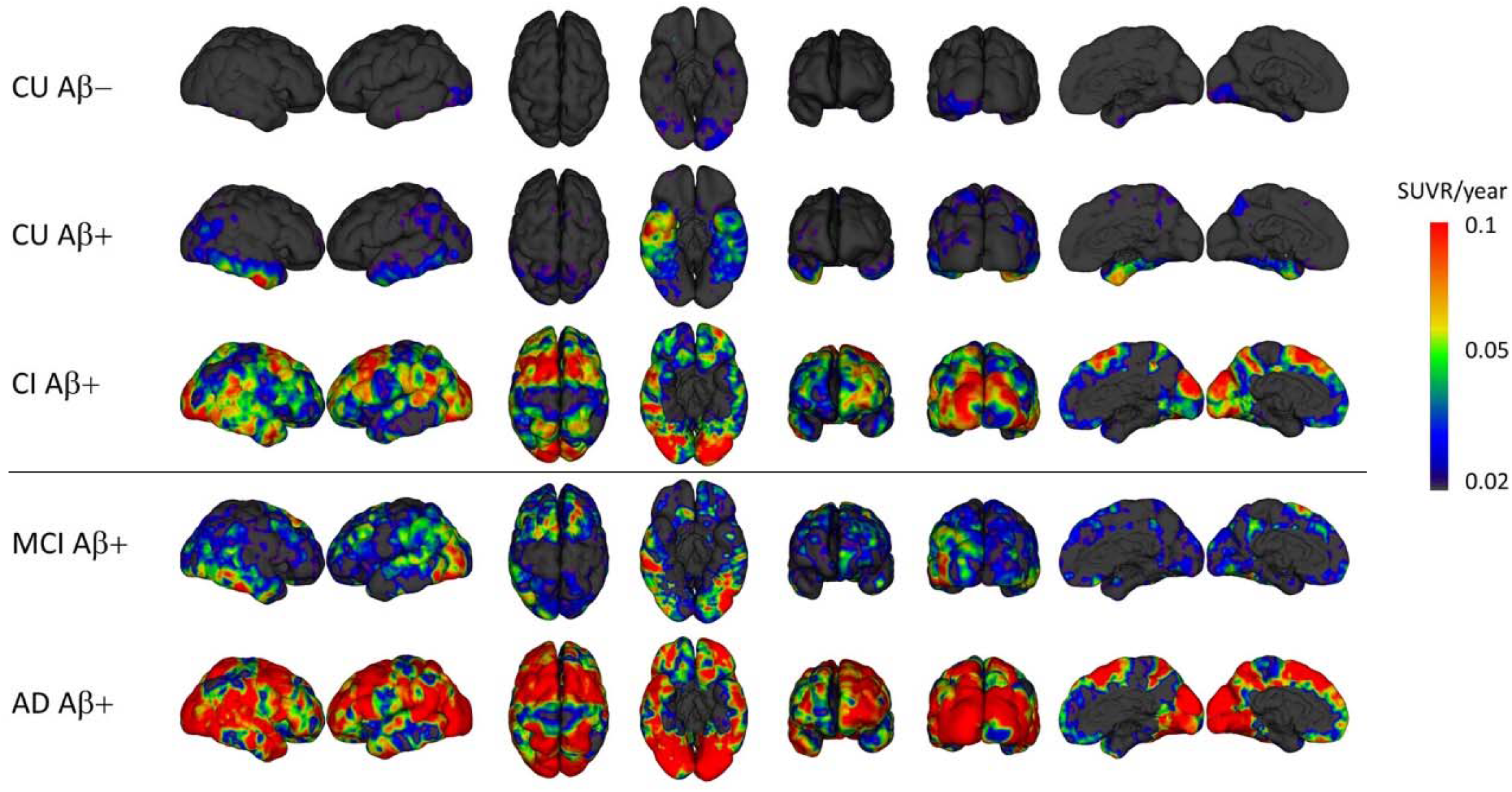
Annual rate of ^18^F-MK6240 accumulation across the clinical groups (cerebellar cortex reference region) Vertex-based surface analysis demonstrating the spatial distribution and rate of ^18^F-MK6240 accumulation (SUVR/ year) for each clinical group, normalized to the cerebellar cortex. Abbreviations: CU = cognitively unimpaired; CI = cognitively impaired; MCI = mild cognitive impairment; and AD = Alzheimer’s disease dementia.

### Tau accumulation in composite brain regions of interest

Baseline SUVR, annual tau SUVR change, and annual tau SUVR percentage change were estimated in each composite ROI for the clinical groups. Results are shown for two reference regions: a) cerebellar cortex (Table 2, Figure 2A, and Figure 3A); and b) eroded subcortical white matter (Table 3, Figure 2B, and Figure 3B). Tau accumulation across individual brain regions for each of the clinical groups is shown in Figure 4. Scans normalized to the cerebellar cortex reference region showed that CU Aβ-individuals had low levels of tau accumulation in Me (1.1%/ year) and MT (1.3%/ year), and no tau accumulation in Te and R (<1%/ year) (Supplementary Table 1). CU Aβ+ individuals showed tau accumulation in Me (2.8%/ year), MT (3.2%/ year), and Te (2.3%/ year), and very low rates of accumulation in R (0.6%/year) (Supplementary Table 1). MCI Aβ+ individuals showed the highest rates of tau accumulation in MT (1.6%/year) and Te (2.3%/year), with lower rates in Me (0.9%/year) and R (1.3%/year). AD Aβ+ individuals showed the highest rates of tau accumulation in Te (2.3%) and R (3.8%), with a negative rate of change in Me (−1.4%/yr) and lower rate of change in MT (0.6%/year respectively) (Supplementary Table 1).

**Table 2.**
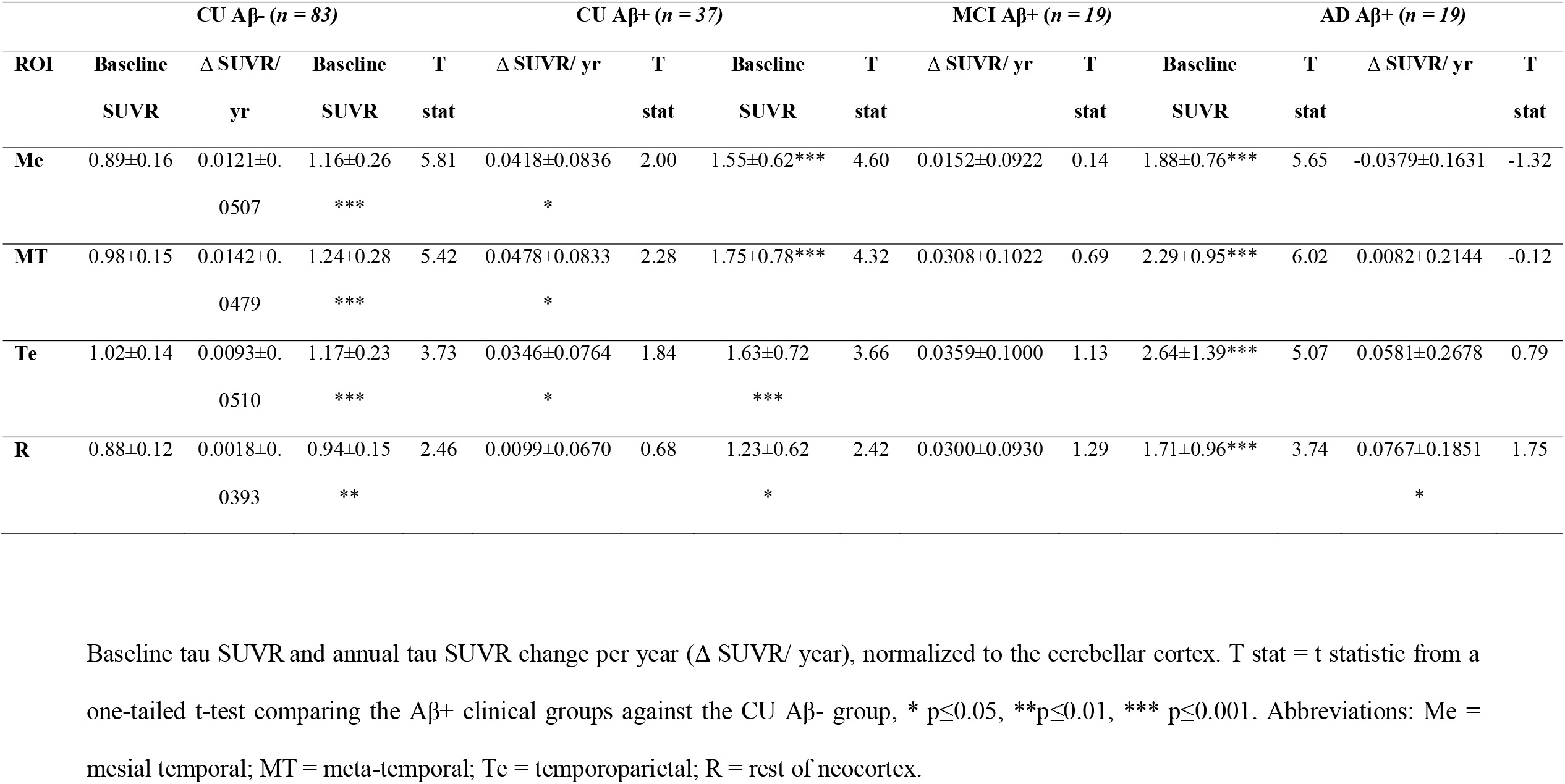
Baseline and annual tau SUVR change (cerebellar cortex reference region)

**Table 3.** Baseline and annual tau SUVR change (eroded subcortical white matter reference region)

| | CU Aβ- (*n* = 83) | | | CU Aβ+ (*n* = 37) | | | MCI Aβ+ (*n* = 19) | | | | AD Aβ+ (*n* = 19) | | | |
| --- | --- | --- | --- | --- | --- | --- | --- | --- | --- | --- | --- | --- | --- | --- |
| ROI | Baseline | $\Delta$ SUVR/ yr | Baseline | T | $\Delta$ SUVR/ yr | T | Baseline | T | $\Delta$ SUVR/ yr | T | Baseline | T | $\Delta$ SUVR/ yr | T |
|  | SUVR |  | SUVR | stat |  | stat | SUVR | stat |  | stat | SUVR | stat |  | stat |
| **Me** | 1.18±0.13 | 0.0233±0.0766 | 1.51±0.36* | 5.41 | 0.0564±0.0938* | 2.04 | 1.88±0.67* | 4.55 | 0.0245±0.1386 | 0.04 | 2.07±0.55** | 7.07 | -0.0501±0.1141 | -2.67 |
|  |  |  | ** |  |  |  | ** |  |  |  | * |  | ** |  |
| **MT** | 1.30±0.13 | 0.0287±0.0775 | 1.61±0.37* | 5.02 | 0.0646±0.0839* | 2.29 | 2.10±0.75* | 4.61 | 0.0457±0.1342 | 0.53 | 2.51±0.68** | 7.78 | -0.0014±0.1333 | -0.95 |
|  |  |  | ** |  |  |  | ** |  |  |  | * |  |  |  |
| **Te** | 1.36±0.16 | 0.0240±0.0861 | 1.52±0.31* | 2.94 | 0.0440±0.0769 | 1.22 | 1.96±0.66* | 3.90 | 0.0633±0.1049* | 1.72 | 2.83±1.04** | 6.15 | 0.0430±0.1538 | 0.52 |
|  |  |  | * |  |  |  | ** |  |  |  | * |  |  |  |
| **R** | 1.18±0.16 | 0.0131±0.0770 | 1.22±0.21 | 1.21 | 0.0112±0.0798 | -0.12 | 1.47±0.55* | 2.23 | 0.0419±0.0623 | 1.52 | 1.84±0.66** | 4.33 | 0.0707±0.1416 | 1.71 |
|  |  |  |  |  |  |  |  |  |  |  | * |  |  |  |

Baseline tau SUVR and annual tau SUVR change per year ( $\Delta$ SUVR/ yr), normalized to the eroded subcortical white matter reference region. T stat = t statistic from a one-tailed t-test comparing the Aβ + clinical groups against the CU Aβ - group, \* $p \leq 0.05$ , \*\* $p \leq 0.01$ , \*\*\* $p \leq 0.001$ . Abbreviations: Me = mesial temporal; MT = meta-temporal; Te = temporoparietal; R = rest of neocortex.

**Figure 2.**
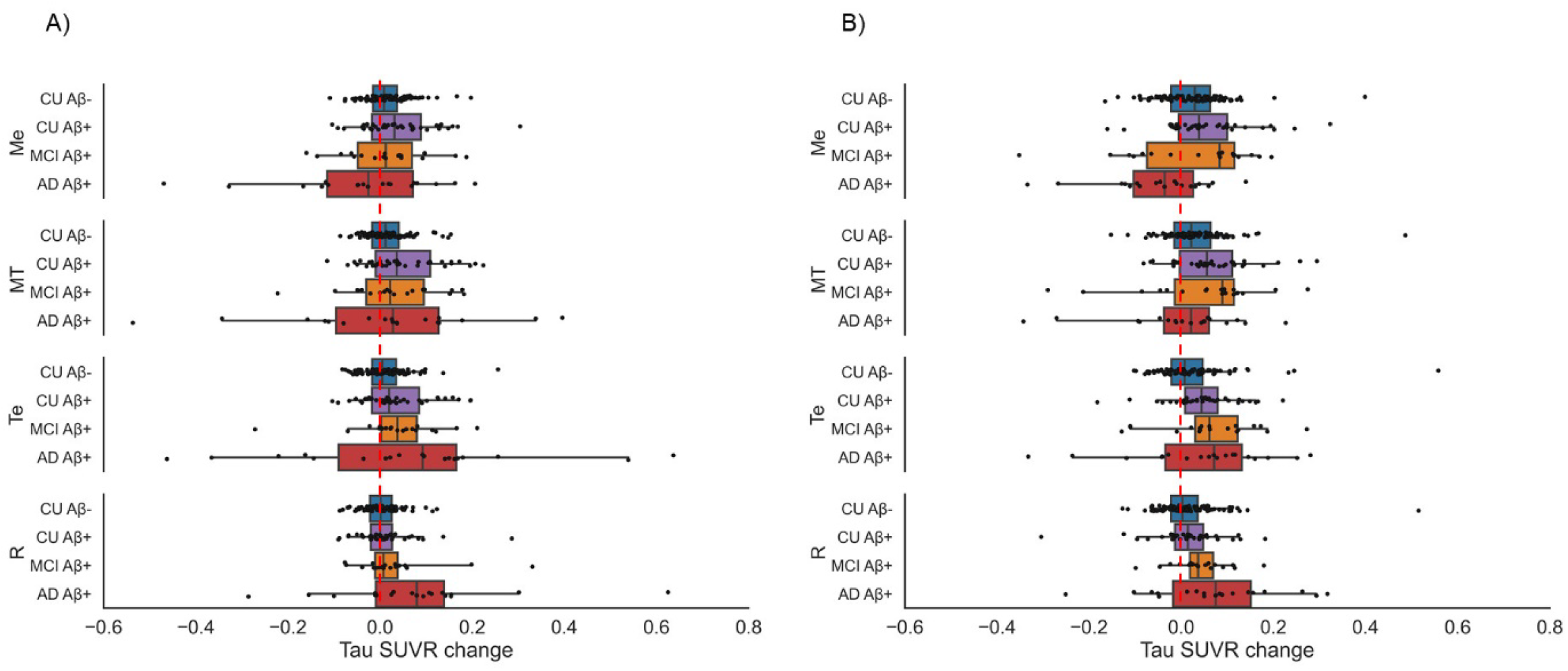
Annual rate of tau SUVR change in composite ROI. Boxplots showing annual tau SUVR change in composite ROI for the clinical groups, normalized to A) cerebellar cortex reference region; and B) eroded subcortical white matter reference region. The red dashed vertical line represents zero change. Within the boxes, the line represents the median value. The whiskers extend from the 5^th^ to the 95^th^ percentile. Abbreviations: CU = cognitively unimpaired; MCI = mild cognitive impairment; AD = Alzheimer’s disease dementia.

**Figure 3.**
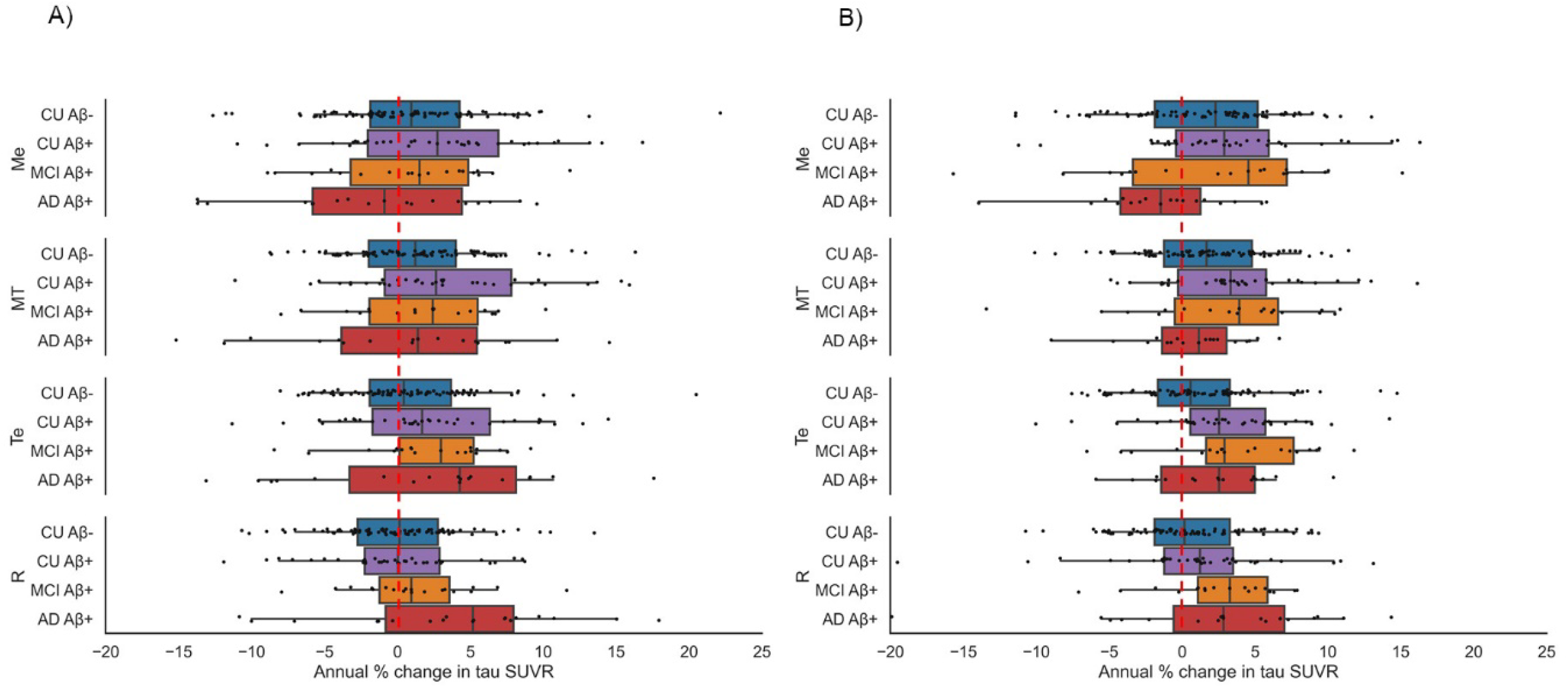
Annual percentage change in tau SUVR in composite ROI. Boxplots showing annual percentage change in tau SUVR in composite ROI for the clinical groups, normalized to A) cerebellar cortex reference region; and B) eroded subcortical white matter reference region. The red dashed vertical line represents zero change. Within the boxes, the line represents the median value. The whiskers extend from the 5^th^ to the 95^th^ percentile. Abbreviations: CU = cognitively unimpaired; MCI = mild cognitive impairment; AD = Alzheimer’s disease dementia.

**Figure 4.**
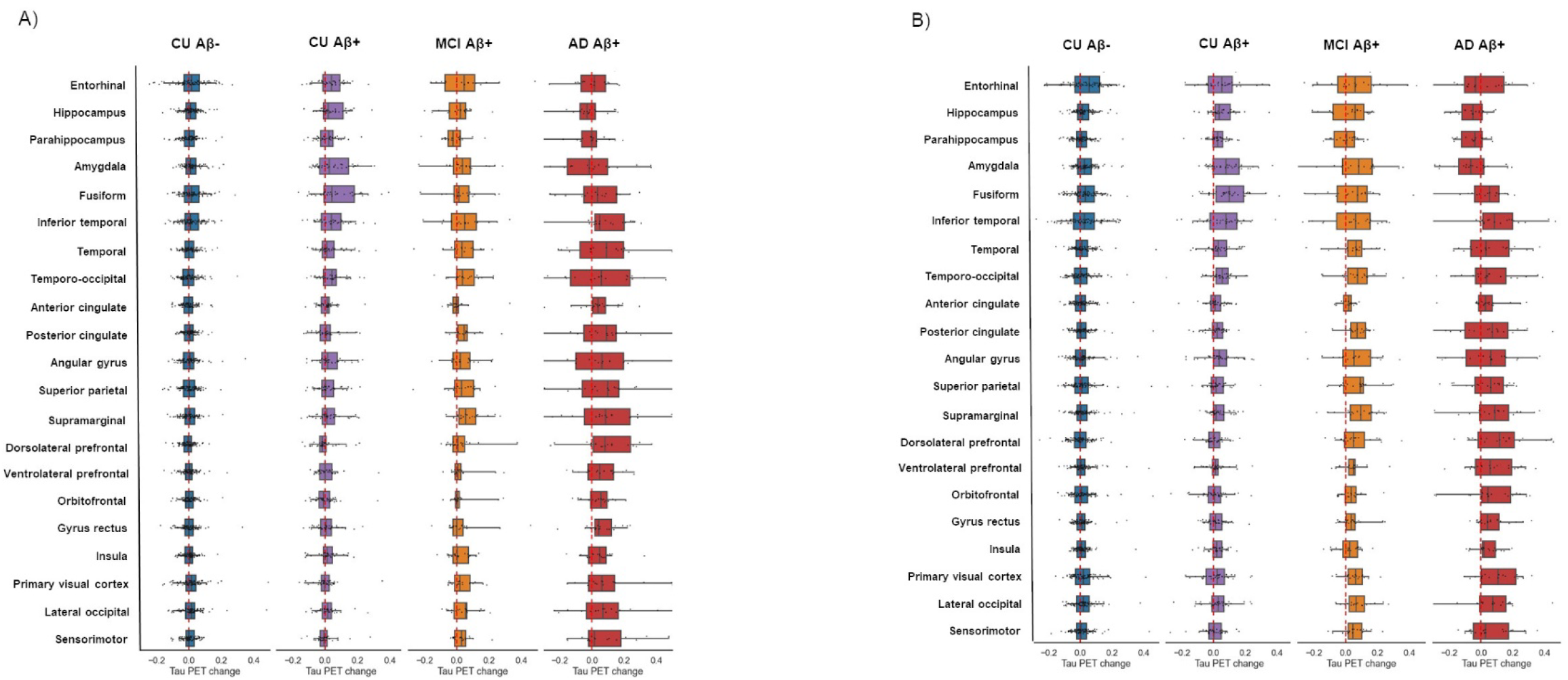
Annual tau SUVR change by region. Annual tau SUVR change across brain regions for the clinical groups, normalized to A) cerebellar cortex reference region; and B) eroded subcortical white matter reference region. The red dashed vertical lines represent zero change. Within the boxes, the line represents the median value. The whiskers extend from the 5^th^ to the 95^th^ percentile. Abbreviations: Me = mesial temporal; MT = meta-temporal; Te = temporoparietal; R = rest of neocortex.

Scans normalized to the eroded subcortical white matter reference region showed that CU Aβ-individuals had low levels of tau accumulation in Me (1.8%/ year), MT (2%/year), but also Te (1.5%/year) (Supplementary Table 2). CU Aβ+ individuals showed tau accumulation in Me (3.2%/ year), MT (3.6%/year), and Te (2.7%/ year), with low rates of accumulation in R (0.97%/year) (Supplementary Table 2). MCI Aβ+ individuals showed tau accumulation in Me (2.4%/ year), and across the cortex (3.1%/year in MT, 3.7%/ year in Te, and 2.7%/year in R) (Supplementary Table 2). AD Aβ+ individuals showed tau accumulation in Te (1.1%/year) and R (2.7%/year), with a negative rate of change in Me (−2.5%/year) and MT (−0.5%/year) (Supplementary Table 2). Higher annual percentage change estimates were observed using the subcortical white matter reference region for the CU Aβ-, CU Aβ+ and MCI Aβ+ groups, while higher estimates were observed using the cerebellar cortex reference region for the AD Aβ+ group.

The effect size for annual percentage change in tau SUVR for the CU Aβ+ group compared to the CU Aβ-group was similar using the SUVR_Cb_ and SUVR_WM_ (Me, *d* = 0.30 vs 0.24; MT, *d* = 0.36 vs 0.31; Te, *d* = 0.30 vs 0.23; R, *d* = 0.09 vs 0.07) (Supplementary Tables 1 and 2). The effect size comparing the annual percentage change in tau SUVR for the MCI Aβ+ group compared to the CU Aβ-group was greater for SUVR_WM_ than SUVR_Cb_ in all composite ROI (Me, *d* = 0.08 vs 0.02; MT, *d =* 0.18 vs 0.08; Te, *d* = 0.41 vs 0.34; R, *d* = 0.38 vs 0.28) (Supplementary Tables 1 and 2). The effect size for annual percentage change in tau SUVR in Te and R was greater for SUVR_Cb_ than SUVR_WM_ when comparing AD Aβ+ to the CU Aβ-group (Te, *d* = 0.23 vs 0.06; R, *d* = 0.60 vs 0.27) (Supplementary Tables 1 and 2). CU Aβ-had higher annual percentage change in tau SUVR than AD Aβ+ in Me and MT, where AD Aβ+ had negative rates of change. The effect size for this difference was greater for SUVR_WM_ than SUVR_Cb_ (Me, *d* = 0.69 vs 0.38; MT, *d* = 0.41 vs 0.10).

### Change in reference region SUV

The rate of SUV change per year in the cerebellar cortex (Supplementary Figure 2A) and eroded subcortical white matter reference region (Supplementary Figure 2B) was not significantly different when comparing CU Aβ+ and MCI Aβ+ to the CU Aβ-group. AD Aβ+ had significantly higher rate of SUV change in the cerebellar cortex (p<0.05) and the eroded subcortical white matter reference regions (p<0.001) than the CU Aβ-group (Supplementary Figure 2). Supplementary Figure 3 shows plots of SUV change by age for the two reference regions. A subset of AD Aβ+ participants under the age of 65 were observed to have an increase in SUV in the eroded subcortical white matter reference region; a finding that was not observed for the cerebellar cortex reference region.

### Age, baseline tau and the spatiotemporal trajectory of tau accumulation

The relationship between tau accumulation and age is shown in Figure 5, while the relationship between baseline tau and tau accumulation in composite ROI is shown in Figure 6. The observed spatiotemporal trajectory of tau accumulation (as normalized to the cerebellar cortex) is shown in Figure 7.

**Figure 5.**
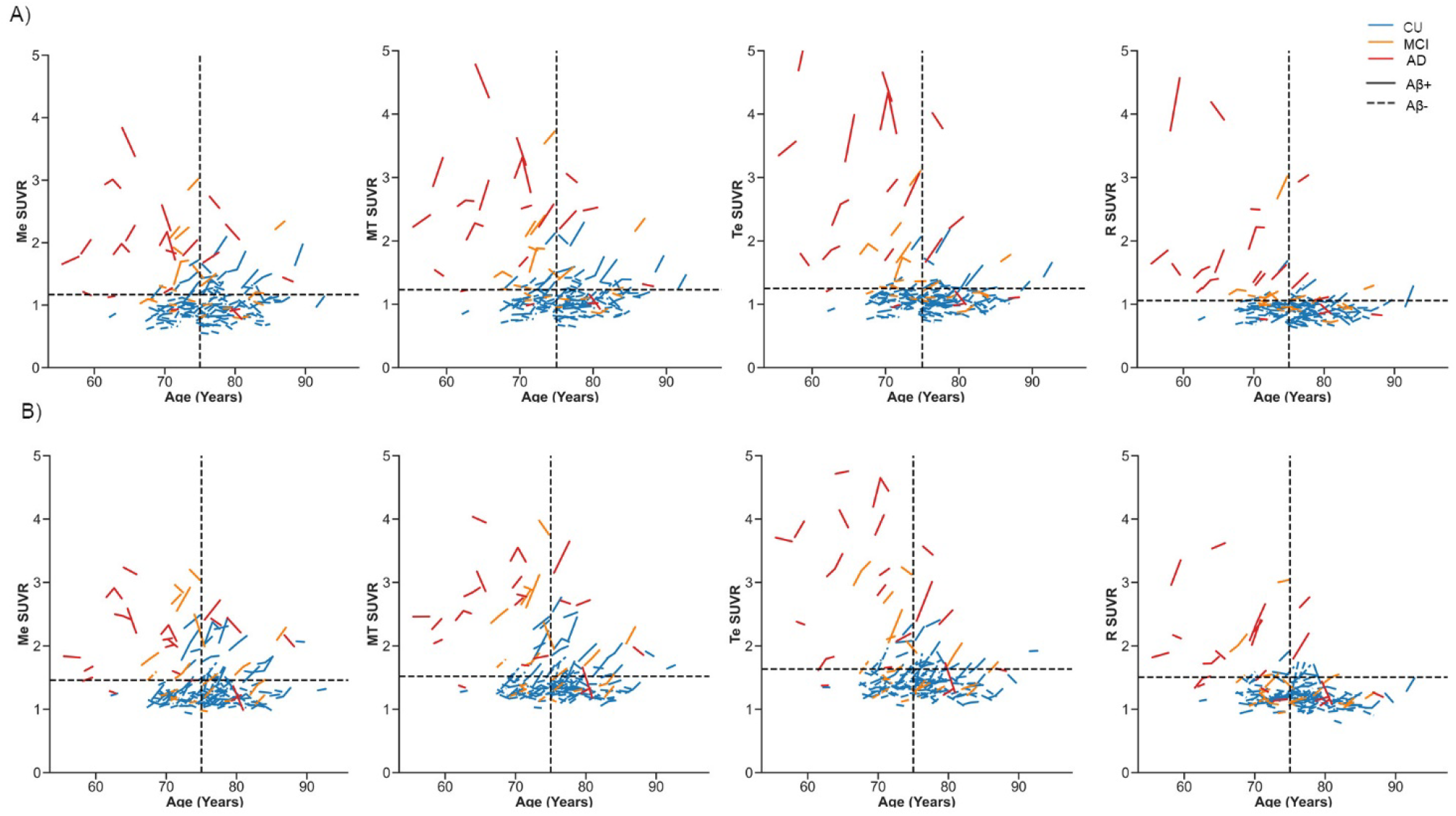
Tau change in composite ROI by age. Tau SUVR in composite ROI by age, normalized to A) cerebellar cortex reference region; and B) eroded subcortical white matter reference region. The vertical black dashed line separates participants above and below 75 years of age. The horizontal black dashed line represents the 95% percentile of CU Aβ-. Blue = cognitively unimpaired (CU). Orange = MCI. Red = AD dementia. Dash lines represent Aβ- and solid lines represent Aβ+. Abbreviations: Me = mesial temporal, MT = meta-temporal, Te= temporoparietal, and R = rest of neocortex.

**Figure 6.**
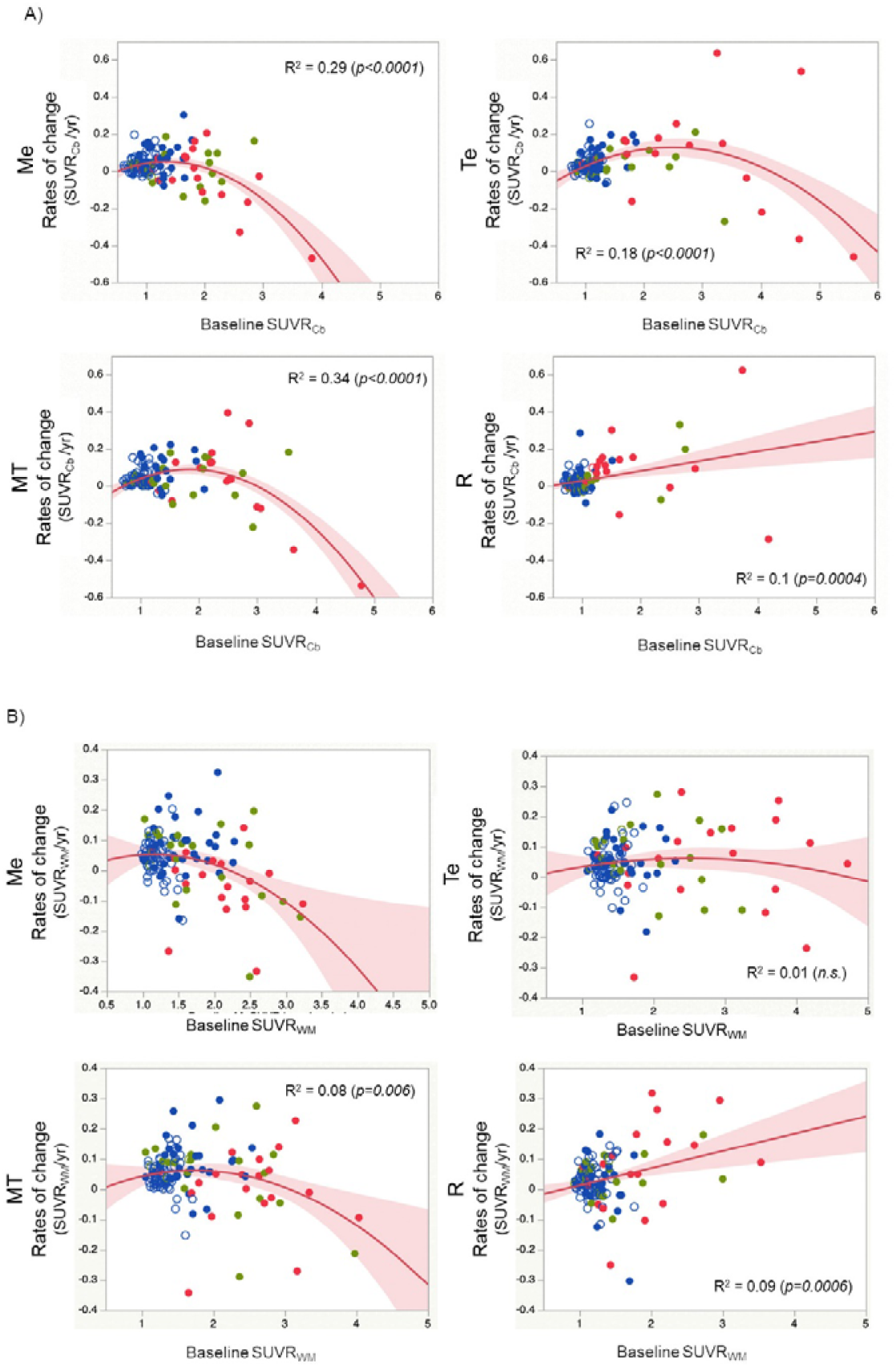
Relationship between baseline tau and rate of accumulation in composite ROI. Relationship between baseline tau SUVR and rate of change (SUVR/ year) for each composite ROI, normalized to A) cerebellar cortex reference region; and B) eroded subcortical white matter reference region. Blue = cognitively unimpaired; green = MCI; red = Alzheimer’s disease dementia; empty circles = Aβ-; solid circles = Aβ+.

**Figure 7.**
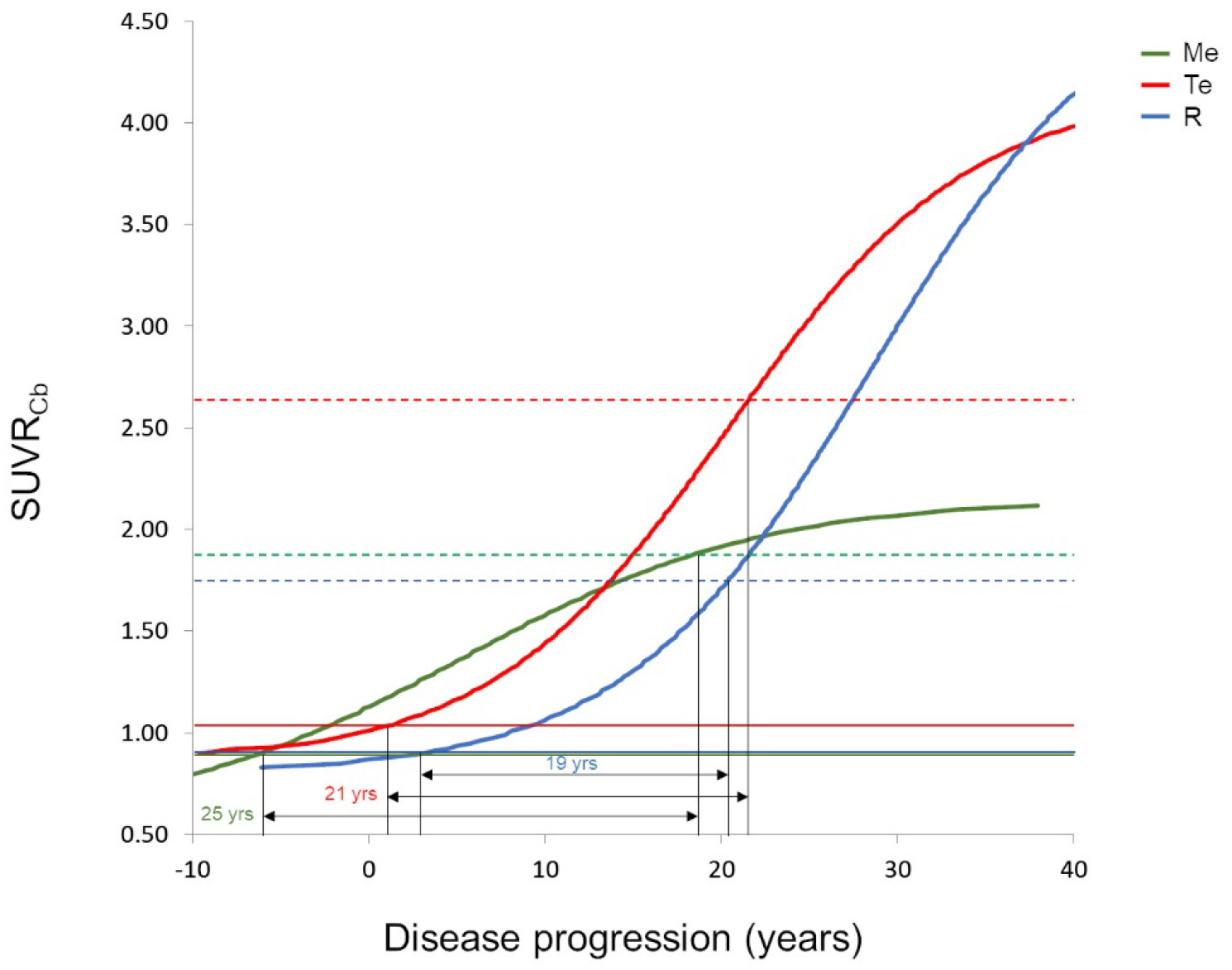
Spatiotemporal trajectory of tau accumulation. Figure 7 shows the natural history of regional tau accumulation measured as ^18^F-MK6240 SUVR normalized to the cerebellar cortex reference region, as interpolated from the data. Solid horizontal lines represent the mean CU Aβ-SUVR_Cb_ values for the composite ROI (Me: 0.89, Te: 1.02; R: 0.88). Dashed horizontal lines represent the mean AD Aβ+ SUVR_Cb_ values for the composite ROI (Me: 1.88, Te: 2.64, R: 1.71). To reach the mean SUVR_Cb_ observed in AD Aβ+ from the mean SUVR_Cb_ observed in CU Aβ-, it takes approximately 25 years in Me, 21 years in Te, and 19 years in R. Abbreviations: Me = mesial temporal (green), Te= temporoparietal (red), and R = rest of neocortex (blue).

## Discussion

Serial tau imaging with ^18^F-MK6240 was able to detect tau accumulation in both cognitively unimpaired (CU) and cognitively impaired participants (MCI and AD in equal proportion) who were Aβ+ at baseline. Tau accumulation rates were evaluated independently using composite ROI and vertex-based surface analyses, with concordant results for these groups. Accumulation of tau was predominantly observed in mesial temporal and temporoparietal regions in CU Aβ+, in temporoparietal regions in MCI Aβ+, and in frontal regions in AD Aβ+. ROI and vertex-based analyses showed low rates of tau SUVR_Cb_ accumulation in CU Aβ-, confined to mesial and meta-temporal regions, while low rates of tau SUVR_WM_ accumulation were also observed to involve temporoparietal regions.

Annual rates of tau SUVR_WM_ accumulation were higher for CU Aβ-, CU Aβ+, and MCI Aβ+ groups compared to SUVR_Cb_, with fewer observed outliers. However, tau SUVR_Cb_ accumulation rates were higher for AD Aβ+. A subset of young (<65 years of age) AD Aβ+ individuals were observed to have a SUV increase in the eroded subcortical white matter reference region, due to either tau accumulation or spill-over of the ^18^F-MK6240 signal into the eroded subcortical white matter mask for this reference region, which lowered the estimated SUVR_WM_ and the yearly rate of change for these individuals.

As observed with Aβ, the regional rates of tau accumulation were dependent on baseline tau burden. AD Aβ+ with very high levels of baseline tau (SUVR>3) showed either a plateau or decline in tau SUVR. This was most apparent in mesial temporal and temporoparietal regions. Plots of regional tau SUVR and age showed that participants with high baseline tau (SUVR>3) were young (<75 years of age) and cognitively impaired (AD Aβ+ > MCI Aβ+). This study also estimated the spatiotemporal trajectory of tau accumulation. While tau starts accumulating early in Me (reaching the threshold for abnormality ∼6.6 and ∼7.5 years earlier than in Te and R, respectively), it slows down in the Me as the disease progresses. It takes about 25 years to get from the levels of CU Aβ-SUVR to the SUVR observed in AD patients, and about 18 years after crossing the threshold of abnormality. Conversely, tau accumulation in Te accelerates, starting to insinuate a plateau at very high tau levels, while tau starts accumulating later in the R composite region with less tendency to plateau at very high levels. In Te, it takes about 21 years to get from the levels of CU Aβ-SUVR to the SUVR observed in AD patients, and about 15 years after crossing the threshold of abnormality. In R, where tau starts accumulating much later, it takes about 19 years to get from the levels of Aβ-CU SUVR to the SUVR observed in AD patients, and about 10 years after crossing the threshold of abnormality.

Minimal tau accumulation in CU Aβ- is consistent with prior longitudinal analyses (13, 19, 21-23), and consistent with observations that tau accumulation is accelerated in the context of Aβ (8, 19, 24). The low rates of tau accumulation in CU Aβ-confined to mesial temporal and temporal regions may be consistent with primary age-related tauopathy (PART) (25). Aβ+ younger individuals (<65 years) have been observed to have a higher baseline cortical tau burden (21, 26), and more rapid accumulation in longitudinal tau ^18^F-flortaucipir PET analyses (21, 26). Few participants in this study were under 65 years of age (7/38 Aβ+ cognitively impaired participants), which may have impacted the group-level rate of tau accumulation.

Based on the results in this study, there are a few points of relevance to AD clinical trials that opt to use tau ^18^F-MK6240 as an outcome measure. Selection of composite ROI for use in a therapy trial should take into consideration that a meta-temporal composite may be ideal for early detection and useful in the assessment of tau accumulation at the preclinical disease stages, while a neocortical composite focusing on temporoparietal regions may be more sensitive to detect changes at the symptomatic stages of disease. The observation of a plateau or decline in the rate of tau accumulation in AD Aβ+ participants with very high levels of baseline tau should be kept in mind when interpreting results of disease-modifying therapies that are aiming to demonstrate clearance or slowing of tau accumulation. Reference region selection is also another important consideration. While both the cerebellar cortex and an eroded subcortical white matter reference region may be suitable to be used for clinical trials targeting preclinical AD, the cerebellar cortex reference region would be preferred for trials in symptomatic AD (particularly for a cohort recruiting younger participants).

### Limitations

There were a few important limitations to this study. The sample size was relatively small, particularly with low numbers in the Aβ+ cognitively impaired groups. Participants in this study had high levels of education and did not have significant medical or psychiatric comorbidities. AD Aβ+ participants had mild dementia (MMSE 22.2±3.5). While these features may be consistent with characteristics of participants recruited to clinical trials, individuals with more severe disease and medical comorbidities, were not represented in this sample and thus may not be widely generalizable. Additionally, the duration of follow-up in this study was shorter than would typically be expected for a preclinical AD trial. This study was also limited by the lack of a replication cohort to validate these findings.

## Conclusion

Longitudinal tau ^18^F-MK6240 PET detected tau accumulation in both preclinical and symptomatic Alzheimer’s disease with 12-24 months follow-up. The regional rate of tau accumulation was influenced by baseline tau burden. Interpolation of the data suggests that tau accumulation takes approximately two decades to reach the levels observed in AD. The use of tau ^18^F-MK6240 PET as an outcome measure for clinical trials should take into consideration the selection of composite ROI, regional baseline tau burden, and choice of reference region, depending on whether the trial is targeting preclinical or symptomatic disease stages.

## Data Availability

The datasets used and/or analyzed during the current study are available from the corresponding author upon reasonable request.

## Declarations

### Ethics approval and consent

This study was approved by the Austin Health Human Research Ethics Committee and written informed consent was obtained from all participants.

### Consent for publication

All participants gave written consent for publication of de-identified data.

### Author contributions

CCR and VLV contributed to study conception and design. VLV also contributed to generation of the figures highlighting the relationship between baseline tau and accumulation, and the natural history of tau accumulation. NK contributed to statistical and image analysis, generation of figures, interpretation of results, and wrote the manuscript. VD contributed to data processing, analysis, vertex-based surface analysis, and interpretation of results. PB contributed to data processing, analysis, and interpretation of results. All authors critically reviewed the manuscript for intellectual content.

## Acknowledgements

The data used in the preparation of this article was obtained from the Australian Imaging Biomarkers and Lifestyle flagship study of aging (AIBL), funded by the Commonwealth Scientific and Industrial Research Organization (CSIRO), National Health and Medical Research Council (NHMRC), and participating institutions. AIBL researchers are listed at www.aibl.csiro.au. The authors thank all participants who took part in this study, as well as their families.

## Abbreviations

CU: cognitively unimpaired
Me: mesial temporal composite
MT: meta-temporal composite
Te: temporoparietal composite
R: rest of neocortex composite
SUVR: standardized uptake value ratio
SUVR_Cb_: standardized uptake value ratio normalized to a cerebellar cortex reference region
SUVR_WM_: standardized uptake value ratio normalized to an eroded subcortical white matter reference region
MTL: mesial temporal lobe
AIBL: Australian Imaging Biomarkers and Lifestyle Study of Ageing

