## Supplementary Material for "Rates of regional tau accumulation in ageing and across the Alzheimer’s disease continuum: An AIBL ^18^F-MK6240 PET study"

**Supplementary Figure 1 Annual rate of $^{18}$F-MK6240 accumulation across the clinical groups (eroded subcortical white matter reference region)**

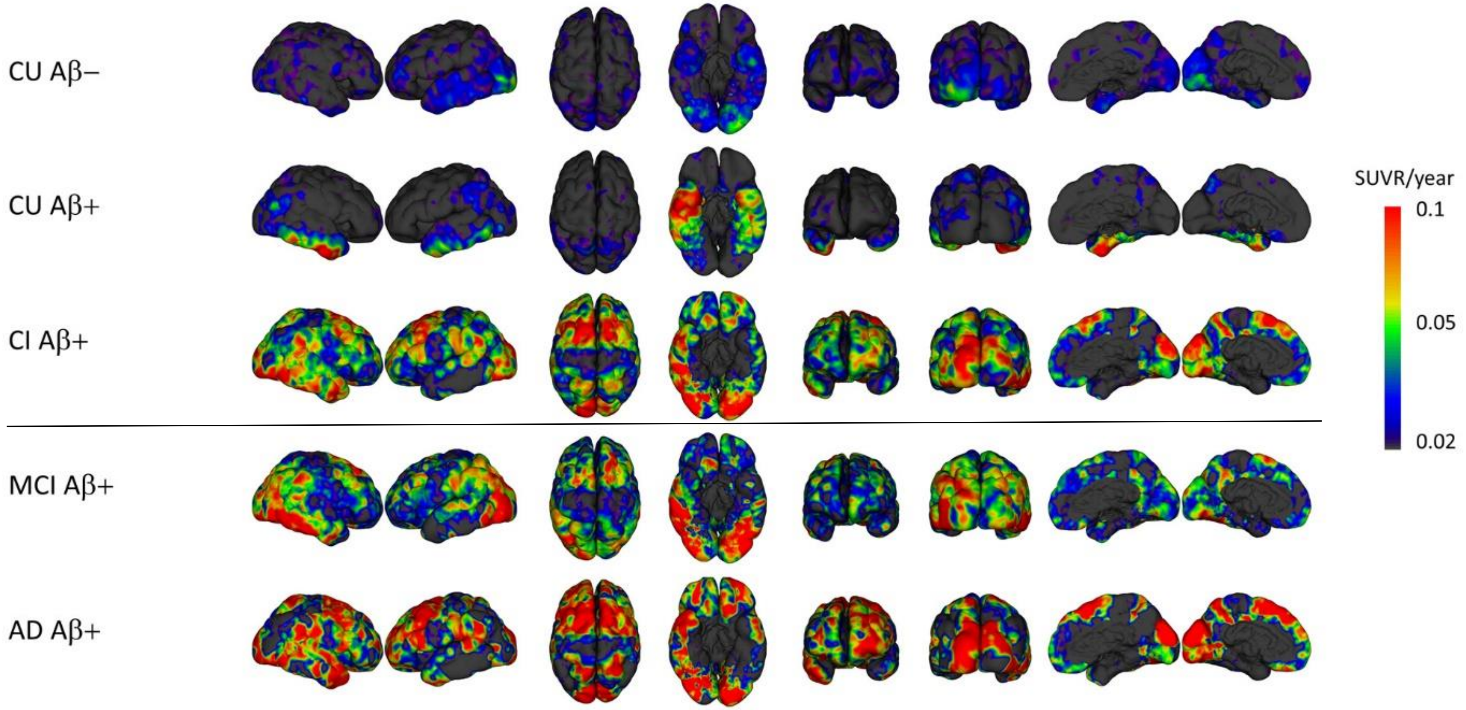


Vertex-based surface analysis demonstrating the spatial distribution and rate of $^{18}$F-MK6240 accumulation (SUVR/ year) for each clinical group, normalized to the eroded subcortical white matter reference region. Abbreviations: CU = cognitively unimpaired; CI = cognitively impaired; MCI = mild cognitive impairment; and AD = Alzheimer's disease dementia.

**Supplementary Table 1 Annual percentage change in tau SUVR (cerebellar cortex reference region)**

| | CU Aβ- (*n = 83)* | CU Aβ+ (*n = 37)* | | | MCI Aβ+ (*n = 19)* | | | AD Aβ+ (*n = 19)* | | |
|---|---|---|---|---|---|---|---|---|---|---|
| **Composite ROI** | **% Δ SUVR/ yr** | **% Δ SUVR/ yr** | **T stat** | ***d*** | **% Δ SUVR/ yr** | **T stat** | ***d*** | **% Δ SUVR/ yr** | **T stat** | ***d*** |
| **Me** | 1.05±5.49 | 2.82±6.37 | 1.55 | 0.30 | 0.93±5.48 | -0.09 | 0.02 | -1.36±7.19 | -1.63 | 0.38 |
| **MT** | 1.25±4.72 | 3.23±6.19* | 1.92 | 0.36 | 1.63±4.84 | 0.31 | 0.08 | 0.62±7.69 | -0.34 | 0.10 |
| **Te** | 0.73±4.65 | 2.31±5.86 | 1.58 | 0.30 | 2.27±4.50 | 1.31 | 0.34 | 2.25±8.08 | 0.79 | 0.23 |
| **R** | 0.04±4.40 | 0.56±6.37 | 0.51 | 0.09 | 1.26±4.37 | 1.09 | 0.28 | 3.81±7.70* | 2.06 | 0.60 |

Annual tau SUVR percentage change per year (% Δ SUVR/ yr). T stat = t statistic from a one-tailed t-test comparing the Aβ+ clinical groups against the CU Aβ- group, * p≤0.05, **p≤0.01, *** p≤0.001. Effect size = Cohen's *d.* Abbreviations: Me = mesial temporal; MT = meta-temporal; Te = temporoparietal; R = rest of neocortex.

**Supplementary Table 2 Annual percentage change in tau SUVR (eroded subcortical white matter reference region)**

| | CU Aβ- (*n = 83)* | CU Aβ+ (*n = 37)* | | | MCI Aβ+ (*n = 19)* | | | AD Aβ+ (*n = 19)* | | |
|---|---|---|---|---|---|---|---|---|---|---|
| **Composite ROI** | **% Δ SUVR/ yr** | **% Δ SUVR/ yr** | **T stat** | ***d*** | **% Δ SUVR/ yr** | **T stat** | ***d*** | **% Δ SUVR/ yr** | **T stat** | ***d*** |
| **Me** | 1.83±5.93 | 3.23±5.84 | 1.20 | 0.24 | 2.38±7.50 | 0.34 | 0.08 | -2.52±6.70** | -2.82 | 0.69 |
| **MT** | 2.03±5.48 | 3.63±4.78 | 1.54 | 0.31 | 3.07±6.18 | 0.73 | 0.18 | -0.53±6.81* | -1.75 | 0.41 |
| **Te** | 1.51±5.75 | 2.71±4.71 | 1.12 | 0.23 | 3.70±4.97 | 1.53 | 0.41 | 1.10±6.87 | -0.27 | 0.06 |
| **R** | 0.82±5.70 | 0.97±5.81 | 0.13 | 0.07 | 2.69±4.00 | 1.35 | 0.38 | 2.68±7.71 | 1.19 | 0.27 |

Annual tau SUVR percentage change per year (% Δ SUVR/ yr). T stat = t statistic from a one-tailed t-test comparing the Aβ+ clinical groups against the CU Aβ- group, * p≤0.05, **p≤0.01, *** p≤0.001. Effect size = Cohen's *d*. Abbreviations: Me = mesial temporal; MT = meta-temporal; Te = temporoparietal; R = rest of neocortex.

**Supplementary Figure 2 Change in reference region (SUV/ year)**

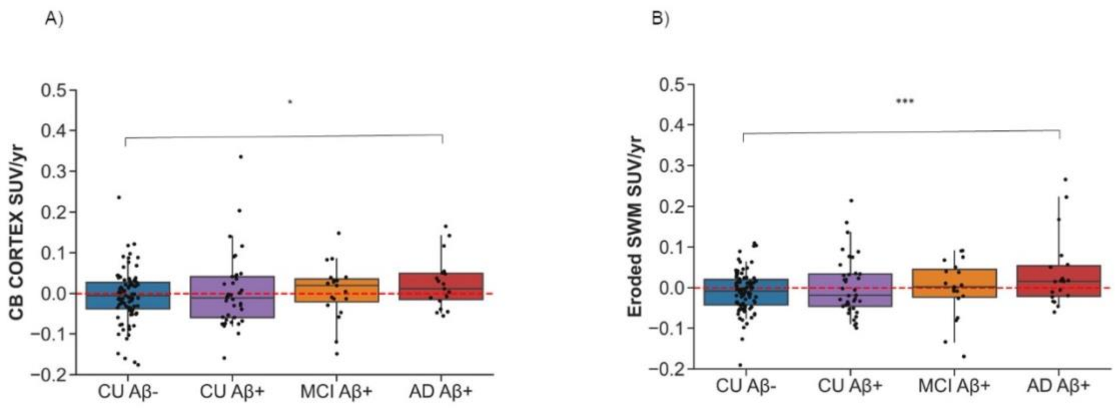


Boxplots showing change (SUV/year) in the A) cerebellar cortex reference region; and B) eroded subcortical white matter reference region, for each of the clinical groups. The red dashed horizontal line represents zero change. The whiskers extend from the 5$^{th}$ to the 95$^{th}$ percentile. *p<0.05, ***p<0.001. Abbreviations: CU = cognitively unimpaired; MCI = mild cognitive impairment; AD = Alzheimer's disease dementia.

**Supplementary Figure 3 Relationship between reference region SUV and age**

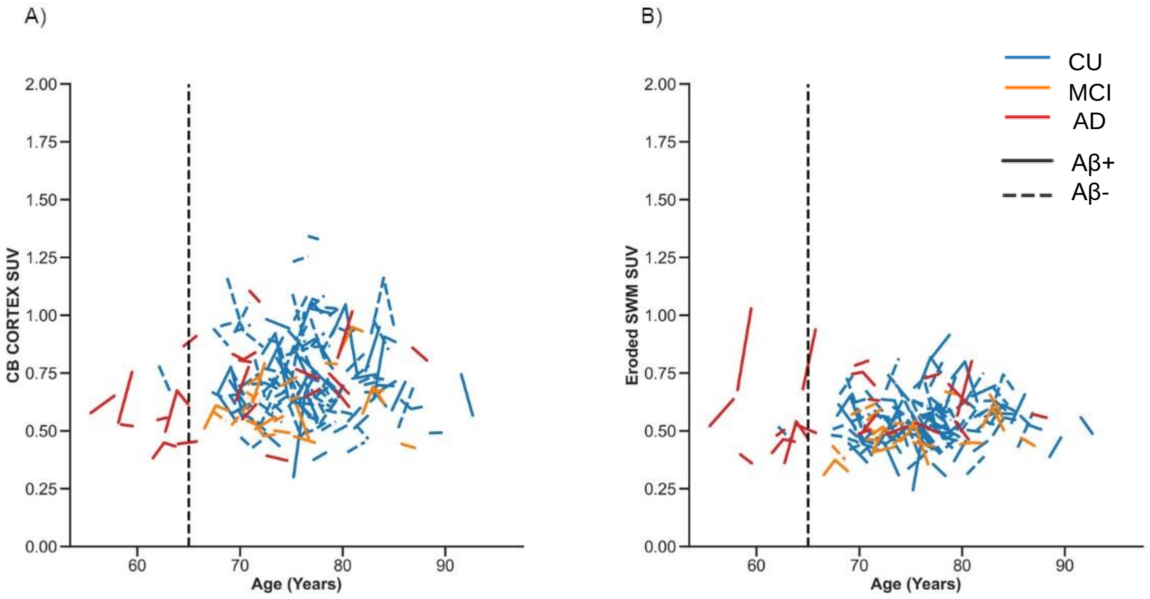


Plots showing standardized uptake value (SUV) for the reference regions A) cerebellar cortex; and B) eroded subcortical white matter versus age. The vertical dashed line separates participants above and below 65 years of age. A subset of AD Aβ+ participants were observed to have an increase in SUV in the eroded subcortical white matter reference region, a change which is not observed for the cerebellar cortex reference region.
